# AI-Empowered Multimodal Data Integration and Knowledge Discovery for Pharmacovigilance

**DOI:** 10.64898/2026.10.20.26365969

**Authors:** Yiming Li, Huiyuan Zhou, Feng-Yu Yeh, Zenan Sun, Jie Zheng, Shayma Alzaidi, Hao Guan, Jianfu Li, Diane L. Seger, Liqin Wang, Manqi Li, Xingxian Li, Taiyang Chen, Joseph M. Plasek, Haifang Li, Aoran Jiang, Pengyu Hong, Fang Li, Wei Zhang, Licong Cui, David W Bates, Li Zhou, Yongqun He, Cui Tao

## Abstract

Biomedical artificial intelligence (AI) increasingly relies on heterogeneous data sources, yet differences in data structure, terminology, evidence quality, and provenance complicate their integration into reliable knowledge resources. Here, we develop an evidence-aware framework for processing and integrating heterogeneous pharmacovigilance data from FDA Adverse Event Reporting System (FAERS) reports, biomedical literature, clinical trials, and electronic health records in Mass General Brigham. The framework combines large language model (LLM)-based information extraction, ontology-driven semantic normalization, cross-source evidence assessment, statistical and machine learning-based prioritization, and ingredient-based adverse drug event (ADE) relationship inference to transform heterogeneous evidence into a structured and evidence-based knowledge resource. The constructed knowledge system contains 11,588 drug nodes, 44,843 ingredient nodes, 3,275 ADE nodes, 993 indication nodes, and more than 2.47 million relationships, with each drug–ADE relationship assigned a confidence score. We further developed a drug-level web interface for accessing the knowledge resource and validated its utility in downstream AI tasks. Specifically, incorporating the developed knowledge resource improved the comprehensiveness, relevance, and overall quality of LLM-based drug-specific ADE synthesis and improved the precision and F1 score of clinical ADE relation extraction. These results demonstrate that AI-assisted processing combined with evidence-aware curation can transform heterogeneous biomedical evidence into reusable, AI-ready knowledge with explicit provenance and evidence strength information. The framework provides a general strategy for integrating heterogeneous biomedical data to support reproducible knowledge construction and downstream biomedical AI applications.

## 1. Introduction

Biomedical literature, clinical records, clinical trials, molecular databases, and observational data provide heterogeneous biological and clinical evidence, represented using different structures, levels of detail, and strength of support [1], [2], [3], [4]. These differences complicate data integration and the construction of knowledge resources that computational models can use reliably[1], [2], [3], [4]. Large language models (LLMs) and other artificial intelligence (AI) methods offer opportunities to extract and synthesize biomedical information at scale [5], but their performance depends on the quality, consistency, and contextual information of the underlying data [6], [7]. However, systematic data processing and harmonization remain challenging, but essential for transforming heterogeneous biomedical information into structured, reusable knowledge and providing a reliable foundation for downstream AI applications in biomedicine.

Pharmacovigilance provides a representative setting for studying these challenges because evidence about adverse drug events (ADEs) is distributed across multiple complementary sources [8]. Spontaneous reporting systems such as the FDA Adverse Event Reporting System (FAERS) provide large-scale postmarketing observations, biomedical publications offer peer-reviewed clinical, biological, and mechanistic evidence, clinical trials record treatment efficacy and side effects under controlled conditions, and electronic health records provide real-world clinical observations [9], [10], [11], [12]. It is evident that information integration across these sources can provide a more comprehensive understanding of drug safety than relying on any individual source, however their differences in how information is structured, described, and supported complicate their synthesis into a coherent and interpretable knowledge representation. Addressing this challenge requires approaches that can extract relevant information, normalize biomedical concepts, reconcile information across sources, and establish sufficient evidence support for relationships incorporated into the structured knowledge resource [1], [2], [3], [4].

Previous studies have established different approaches to constructing knowledge graphs for ADE research. ADEpedia integrates drug product labels and standardized drug and clinical terminologies through XML-to-RDF transformation[13], ontology-based normalization, and a semantic web infrastructure that supports human review and curation. GA-ADE constructs a graph from candidate ADE relationships extracted from biomedical literature, then applies graph algorithms to identify core vertices, cluster related entities, and estimate drug–event associations [14]. DKADE uses drug instructions, pharmacopeia, and ontology-based modeling to construct a drug knowledge graph, extracting entities, relationships, and attributes before applying knowledge fusion and inference to verify and extend the graph [15]. However, the information integration from heterogeneous sources and establishing the reliability and downstream utility of the resulting knowledge graph are not fully investigated and remain challenging because each source presents distinct limitations. Biomedical literature, clinical trials, and electronic health records provide clinically relevant evidence, but extracting drug–ADE relationships from unstructured text and interpreting them in context remain difficult for automated AI methods, including LLMs [16], [17], [18]. In contrast, FAERS provides structured reports that facilitate computational analysis, but its spontaneous reports are subject to underreporting, reporting bias, and data quality issues, and reported drug–ADE associations do not necessarily establish causality [19], [20].

Integrating these sources therefore requires more than data aggregation. Moreover, the utility of the constructed knowledge graph must be demonstrated through downstream AI tasks rather than inferred from its size or coverage alone.

This study therefore develops an AI-empowered evidence-aware framework for processing and integrating heterogeneous biomedical evidence with pharmacovigilance as a use case, and supports AI applications with novel knowledge discovery. The framework (Figure 1) combines evidence-aware knowledge acquisition, semantic normalization, computational prioritization and reasoning, and ingredient-based ADE inference to transform distributed biomedical information into structured and reusable knowledge for AI systems. We validate the developed knowledge resource in two downstream AI applications: 1) graph-to-text generation for medication-specific ADE synthesis based on retrieval-augmented generation (RAG) and 2) knowledge graph (KG)-based ADE relation extraction from clinical notes. We further examine its application to ADE characterization involving medications indicated for pyrexia, glucagon-like peptide-1 receptor agonists (GLP-1 RAs) and antidepressant medications.

**Figure 1.**
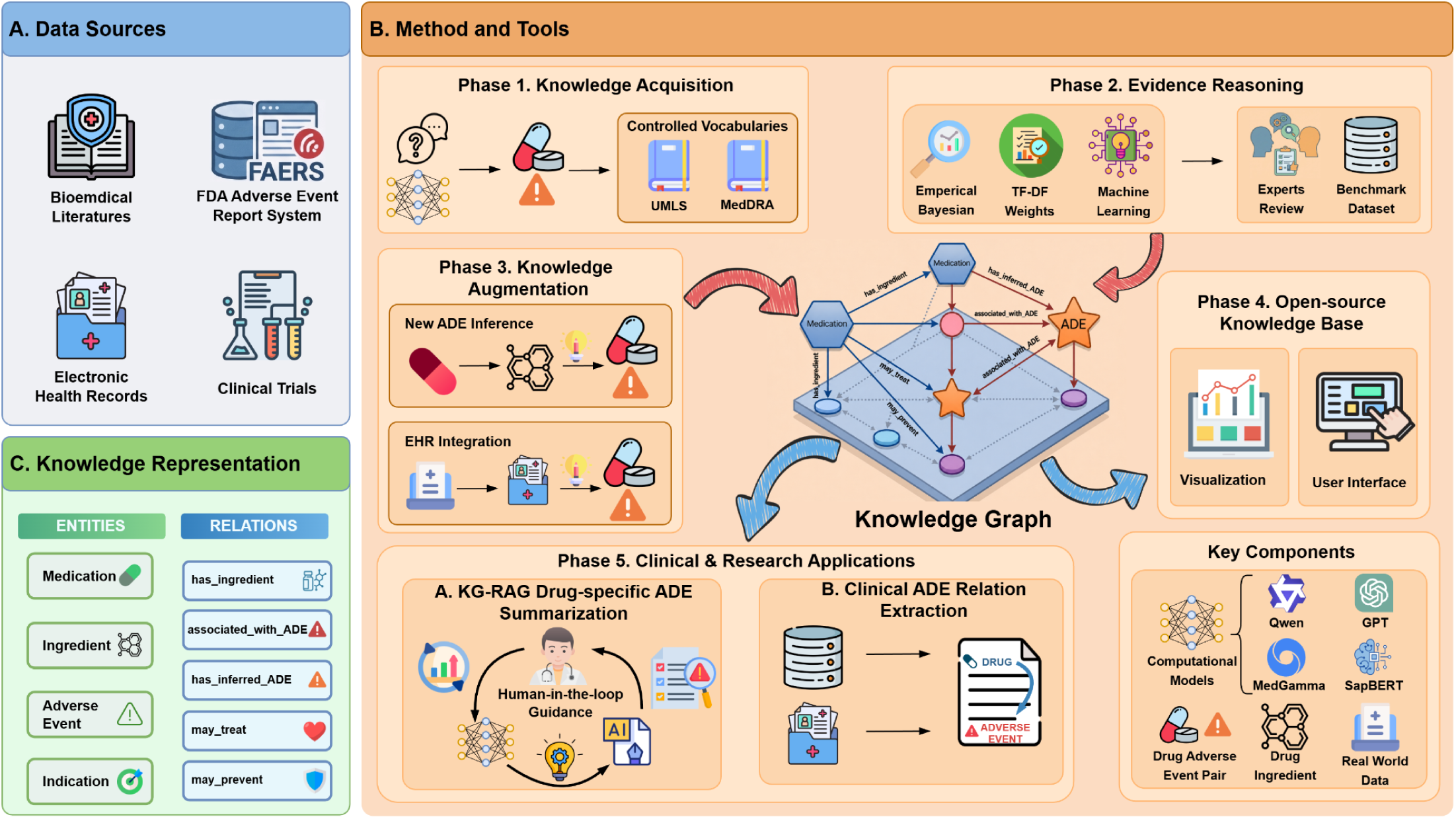
Overview of Study Framework. **Abbreviations:** ***FDA*** The Food and Drug Administration; ***UMLS*** Unified Medical Language System; ***MedDRA*** Medical Dictionary for Regulatory Activities**; *TF-DF*** Term FrequencyF-Document Frequency; ***ADE* Adverse Drug Events; *EHR*** Electronic Health Records; ***KG*** Knowledge Graph; ***RAG*** Retrieval-Augmented Generation; ***GPT*** Generative pretrained transformer

## 2. Methods

### 2.1 Data Sources

We integrated four major data sources (Table 1): spontaneous reports, biomedical literature, clinical trials, and electronic health records (EHRs). Six publicly available benchmark datasets were used to independently evaluate the comprehensiveness of ADE concepts and relations.

**Table 1.**
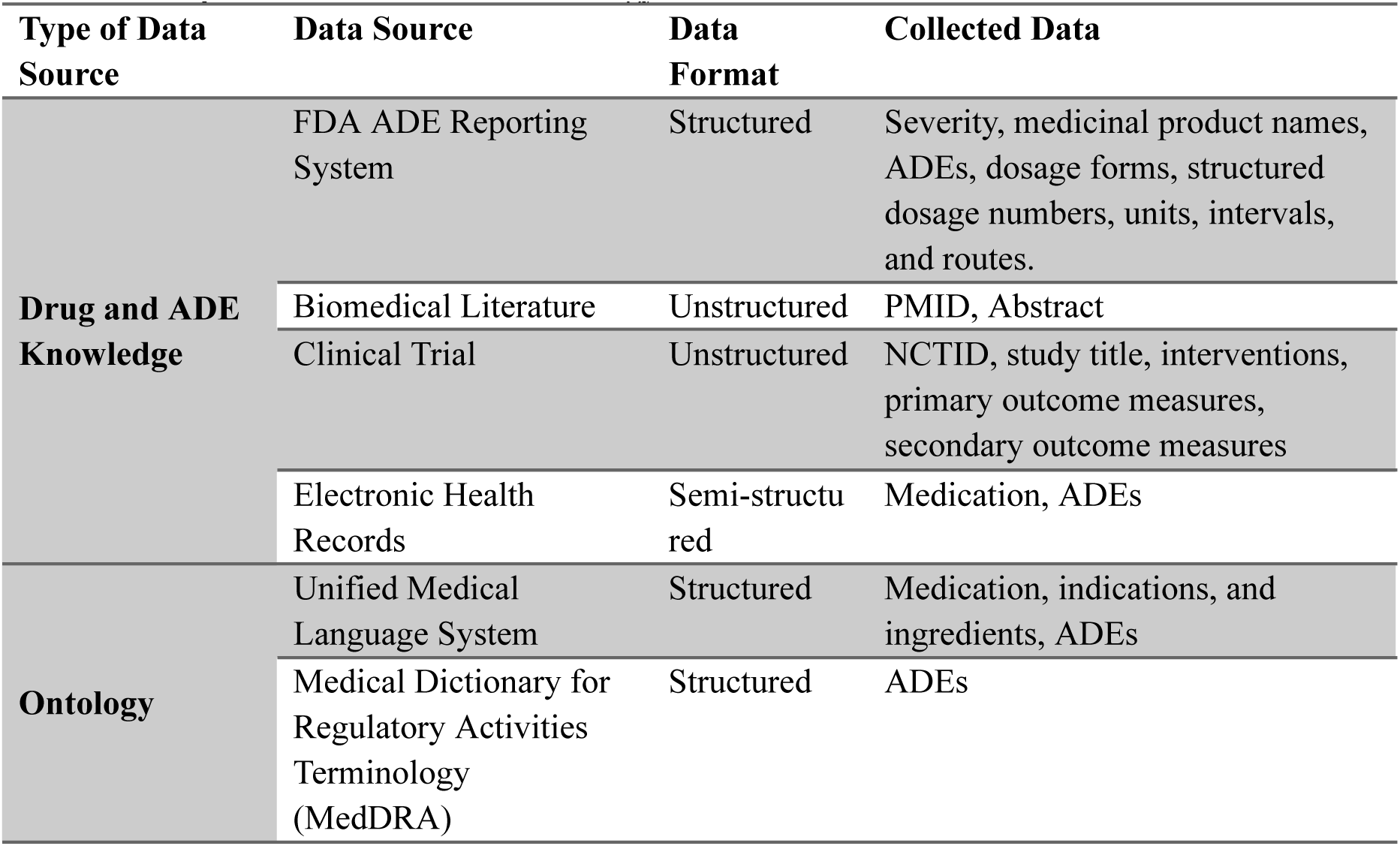
Description of data sources and ontology.

| Type of Data Source | Data Source | Data Format | Collected Data |
| --- | --- | --- | --- |
| **Drug and ADE Knowledge** | FDA ADE Reporting System | Structured | Severity, medicinal product names, ADEs, dosage forms, structured dosage numbers, units, intervals, and routes. |
|  | Biomedical Literature | Unstructured | PMID, Abstract |
|  | Clinical Trial | Unstructured | NCTID, study title, interventions, primary outcome measures, secondary outcome measures |
|  | Electronic Health Records | Semi-structured | Medication, ADEs |
| **Ontology** | Unified Medical Language System | Structured | Medication, indications, and ingredients, ADEs |
|  | Medical Dictionary for Regulatory Activities Terminology (MedDRA) | Structured | ADEs |

#### Spontaneous reports

We collected 21,726,068 reports from the US Food and Drug Administration Adverse Event Reporting System (FAERS) between 2004 and 2024. We extracted reported ADEs, medication product names, dosage forms, dose values, and units, dosing frequency, administration route, and serious clinical outcomes. Serious clinical outcomes included death, life-threatening events, hospitalization, disability, and congenital anomalies.

#### Biomedical literature and clinical trials

We searched PubMed and ClinicalTrials.gov for relevant records through October 17, 2025 using targeted queries related to ADE and adverse drug reactions, which included 151,917 biomedical publications and 76,027 clinical trial reports. For publications, we collected PubMed identifiers (PMIDs) and its abstracts for information extraction. For clinical trials, we collected NCT identifiers (NCTID), titles, intervention information, and primary and secondary outcome measures.

#### Electronic health records

We obtained 4,234,327 documented allergy and adverse reaction records from the Mass General Brigham (MGB) between March 28, 1980 and October 8, 2019 [27], [28], [29]. These records provided observations from routine clinical practice, extending the knowledge base beyond spontaneous reports, publications, and clinical trials.

#### Benchmark datasets

We used six publicly available datasets to evaluate the comprehensiveness: CADEC, MADE1, n2c2, SMM4H 2017, SMM4H 2018, and TAC 2017 [30], [31], [32], [33], [34], [35]. These datasets contain annotated clinical narratives, social media posts, or biomedical texts, with annotations for medication mentions, ADE mentions, or drug-ADE relations. The benchmark datasets were reserved for evaluation and were not used to construct the knowledge graph.

#### Ontology resources

We used the Unified Medical Language System (UMLS) Metathesaurus, version 2025AB, to support biomedical concept normalization and relationship mapping. RxNorm release 2020AA was used to identify medication ingredients and relationships among drug concepts. ADE concepts were standardized using Medical Dictionary for Regulatory Activities (MedDRA) Preferred Terms (PTs).

### 2.2 Knowledge Graph Entities and Relations

The ADE KG (Figure 4a) represents four primary entity types: medications, ingredients, ADEs, and indications. *Medications* represent drug products identified. *Ingredients* indicate their pharmaceutical ingredients and support the identification of related products. *ADEs* represent undesirable clinical outcomes documented in association with medication exposure. *Indications* are the diseases or clinical conditions for which medications are used or investigated.

The KG contains five relationship types. The *associated_with_ADE* relationship links a medication to an ADE identified from the data source. The *has_inferred_ADE* relationship represents a drug-ADE association added through ingredient-based inference. The *has_ingredient* relationship links a medication product to its pharmaceutical ingredient. The *may_treat* relationship links medications to therapeutic indications, and the *may_prevent* relationship connects medications to conditions for which preventive use or effects are represented.

Medication indications were identified using the MRCONSO and MRREL tables of the Unified Medical Language System (UMLS) version 2025AB. We selected English-language preferred terms (ISPREF = ‘Y’) and used UMLS concept identifiers (CUIs) and semantic relations to identify medication-condition associations. The extracted relations were mapped to the *may_treat* or *may_prevent* according to their source relationships and intended meanings.

Medication ingredients were identified using the RXNCONSO and RXNREL files from RxNorm release 2020AA. We examined semantic relationships, including *has_tradename*, *form_of*, and *has_part, to map medication concepts to their corresponding ingredients*. A rule-based regular-expression pipeline removed dose- and strength-specific expressions, such as “50 MG” and “10 ML”, where these expressions were not required to identify the underlying medication concept.

### 2.3 Evidence-aware Knowledge Graph Development

We constructed the ADE KG in Neo4j to represent relationships among drugs, ADEs, and other biomedical entities. Extracted entities were normalized to standardized biomedical concepts to improve consistency across data sources before integration into the graph. ADE concepts were assigned MedDRA identifiers (IDs), and UMLS CUIs were used to support concept linkage across relevant biomedical vocabularies and resources.

Relationships were annotated with its source, calculated confidence score, and supporting metadata, including PubMed ID, FAERS report ID, and NCT ID where applicable. Confidence scores were assigned based on the procedures described below. For medication products, we also recorded whether serious adverse outcomes had been reported in FAERS. This annotation indicates the presence of reported serious outcomes rather than a causal determination of medication-related severity.

To characterize the developed ADE KG, we summarized the distributions of medication and ADE nodes. ADE concept frequencies were also summarized across the integrated sources and visualized using a word cloud (Figure 4c). These summaries describe the scope of ADE information represented in the KG rather than population-level ADE incidence or prevalence.

#### 2.3.1 Evidence Acquisition and Harmonization

The overall workflow for extracting and normalizing information is illustrated in Figure 2a. For biomedical publications, we processed PubMed abstracts. For clinical trials, we aggregated text from relevant metadata, including trial titles, brief summaries, detailed descriptions, interventions, and outcome measures.

**Figure 2.**
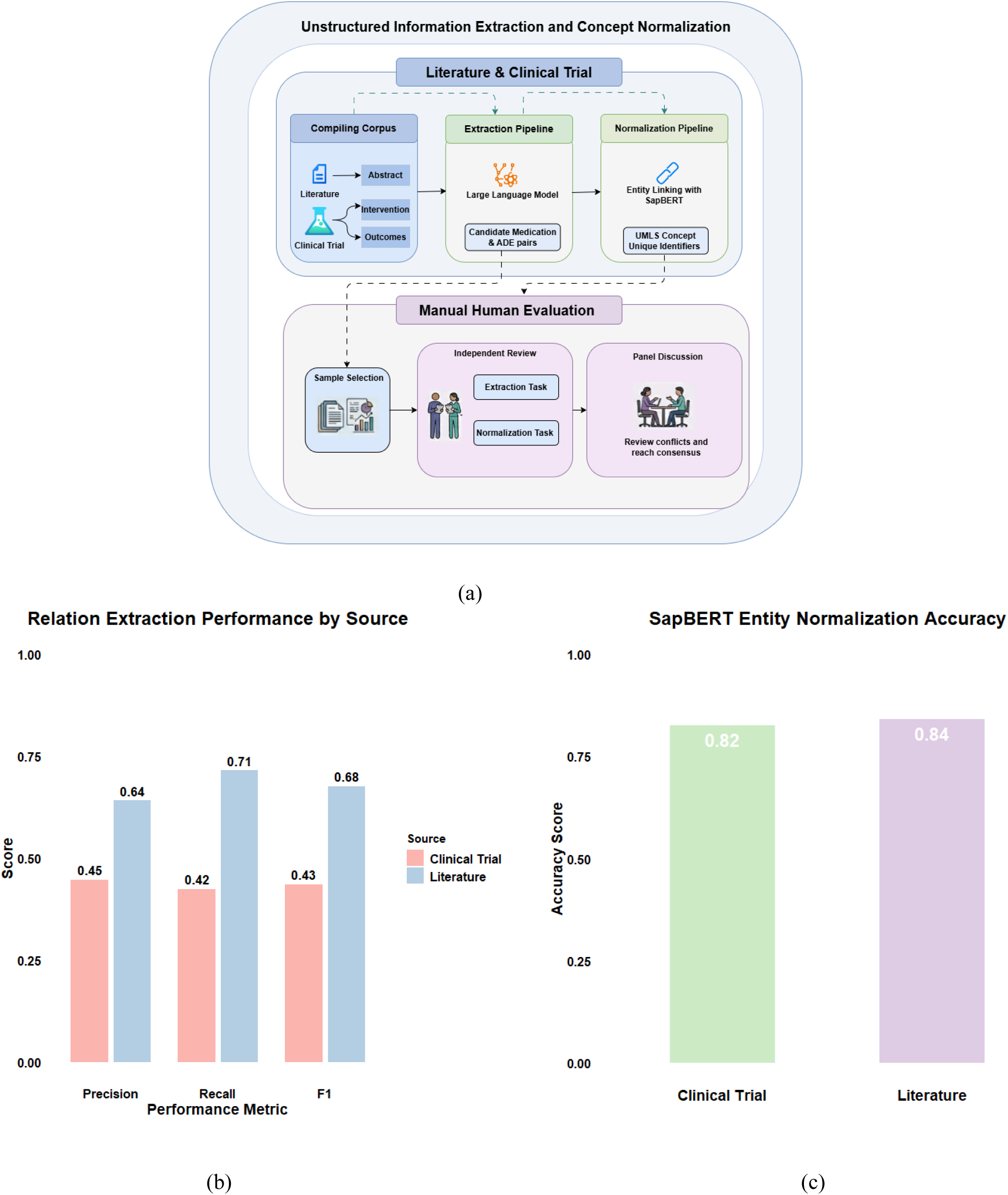
Biomedical information extraction and concept normalization. a) Overview of the extraction and normalization framework b) Evaluation of drug–ADE relation extraction from biomedical literature and clinical trial records c)Evaluation of concept normalization using SapBERT.

We used Qwen3-4B-Instruct-2507 to extract candidate drug-ADE pairs from the biomedical literature and clinical trials using the default parameters. The extraction prompts are provided in the Supplementary Information. Extracted medication and ADE mentions were normalized using SapBERT, a biomedical entity-linking model, to map mentions to UMLS concepts. Medication indications and active ingredients were mapped to standardized concepts where applicable, and ADE concepts were additionally mapped to MedDRA PTs. This process established a consistent representation of drug and ADE concepts across heterogeneous sources.

The normalized relationships were then subjected to the prioritization procedures described in Section 4.3.2. The extraction stage generated candidate relationships; extraction alone did not establish that a relationship was clinically valid or causally attributable to a medication.

#### 2.3.2 Evidence-aware Knowledge Integration and Confidence Estimation

As the availability, completeness, and characteristics of pharmacovigilance evidence differ across data sources, we developed an evidence reasoning framework to prioritize candidate drug–ADE relationships (Fig. 3a). The framework combined cross-source corroboration, empirical Bayesian analysis, term frequency–document frequency weighting, supervised machine learning, and pharmacist review. EHR-derived ADE relationships were excluded from this process because they had been validated by physicians.

**Figure 3.**
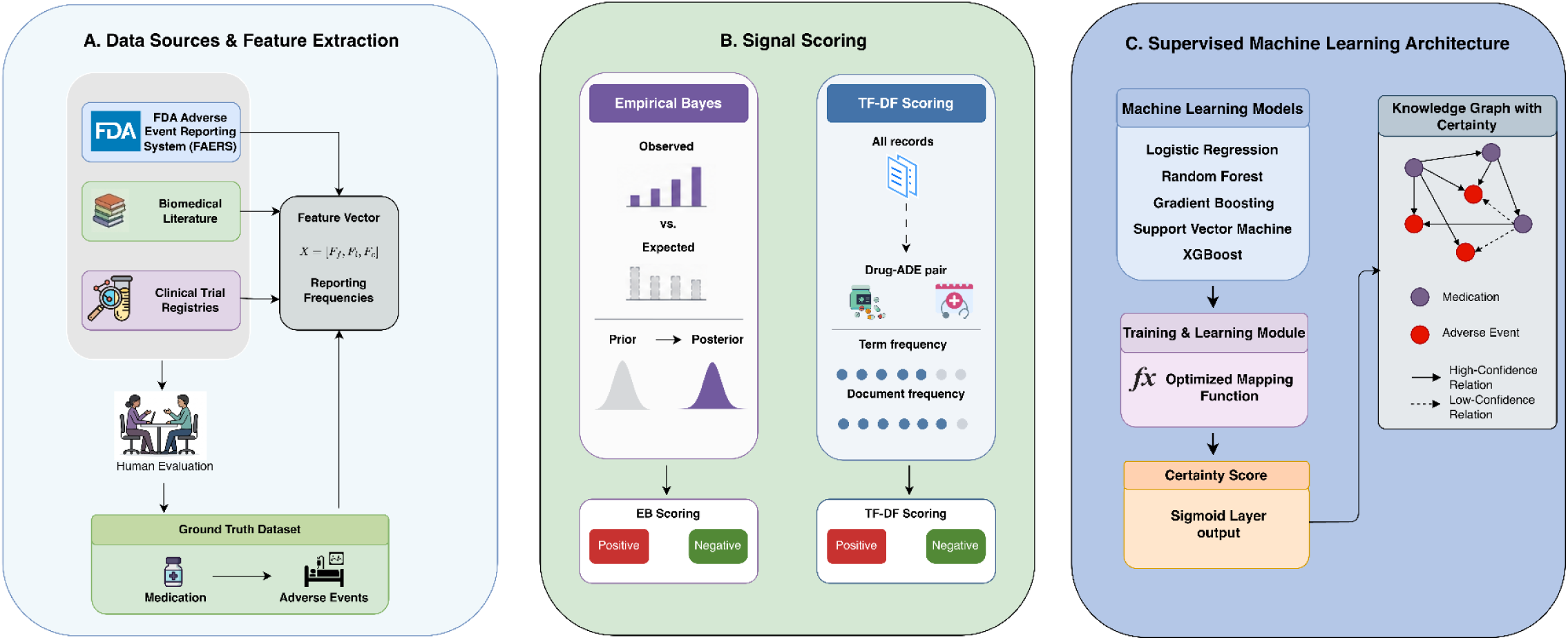
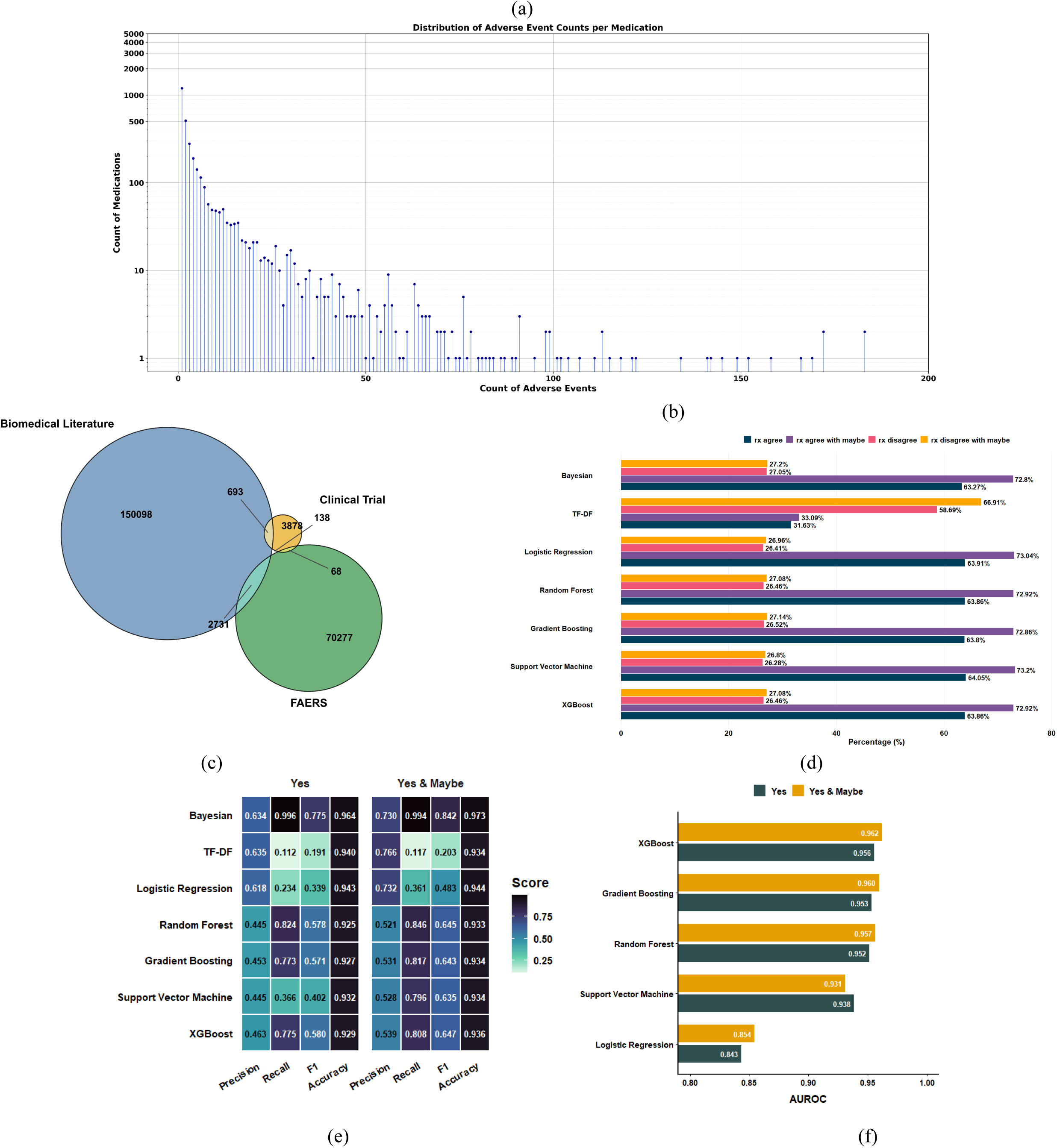
Evidence assessment and prioritization of candidate drug–ADE relationships. (a) Overview of the evidence assessment and prioritization framework. (b) Distribution of the number of ADEs associated with each drug. (c) Venn diagram showing source-specific and overlapping drug–ADE relationships across biomedical literature, clinical trial records, and FAERS. (d) Comparison of computational prioritization results with pharmacist judgments. (e) Precision, recall, and F1-scores for the Poisson–Gamma empirical Bayesian approach, TF–DF, Logistic Regression, Random Forest, Gradient Boosting, SVM, and XGBoost. (f) AUROC values for the supervised classification models under the strict and inclusive pharmacist-label definitions.

##### Cross-source corroboration

Candidate drug–ADE relationships were compared across FAERS, biomedical literature, and clinical trial records. Relationships identified in at least two independent sources were designated as cross-source corroborated. Relationships identified in only one of these sources were subjected to additional statistical or computational prioritization rather than being automatically excluded.

##### Poisson-Gamma Empirical Bayes Analysis

We used a Poisson-Gamma empirical Bayes (EB) model to prioritize drug-ADE pairs identified in only one of the three sources: FAERS, biomedical literature, or clinical trials. Analyses were performed separately for each source. Define *O_d_*_,*a*_ as the observed count of drug-ADE pair (*d*, *a*) occurrences in a given source, *O_d_*_+_ as the total count of drug *d* occurrences across all ADEs, *O*_+*a*_ as the total count for ADE *a* occurrences across all drugs, and *T* the total number of drug-ADE occurrences in the source. Multiple occurrences of the same drug-ADE pair are counted repeatedly. Under a marginal independence model, the expected count was calculated as:

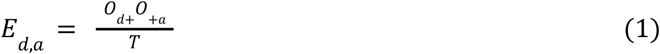

The expected count *E_d_*_,*a*_ provides a source-specific baseline based on the marginal frequencies of the drug and ADE.

We modeled the observed count using a Poisson distribution with a multiplicative enrichment parameter θ*_d_*_,*a*_ :

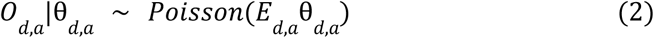

Values of θ*_d_*_,*a*_ = 1 indicate counts consistent with the marginal independence expectation, whereas θ*_d_*_,*a*_ > 1 indicates enrichment relative to that expectation.

As the multiplicative factor θ is strictly greater than 0, we assigned a Gamma prior to the enrichment parameter:

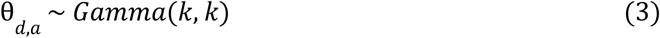

using the shape–rate parameterization. This prior has mean 1 and variance 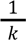. The parameter *k* controls the degree of shrinkage toward the null expectation.

For each source, *k* was estimated by maximizing the marginal likelihood across drug-ADE pairs. Specifically, θ*_d_*_,*a*_ was integrated out of the Poisson–Gamma model, yielding the marginal distribution as shown in Formula (4):

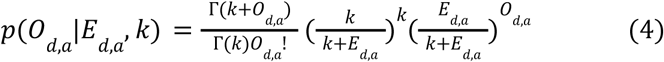

The source-specific estimate *k* was obtained by maximizing the summed log marginal likelihood across all drug-ADE pairs within each source.

Given *k̂*, the posterior distribution of the enrichment parameter for each drug-ADE pair is available in closed form by conjugacy:

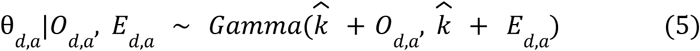

From this posterior distribution, we computed the posterior mean as shown in Formula (6):

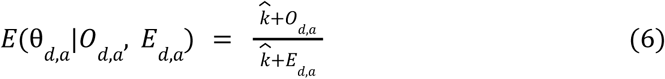

Posterior intervals were summarized using the 5^th^ and 95^th^ percentiles to maintain consistency with the EB05/EB95 convention commonly used in empirical Bayes pharmacovigilance analyses [36]. For relationships identified in only one source, pairs were included if the EB05 exceeded 1, indicating that the lower bound of the posterior credible interval was above the null enrichment value.

##### Term Frequency - Document Frequency- Weights

We developed a term-frequency–document frequency (TF–DF) score to prioritize drug–ADE pairs that lacked cross-source corroboration. Unlike conventional term frequency–inverse document frequency (TF–IDF), which downweights terms occurring in many documents, TF–DF was designed to reward relationships that were both frequently observed and distributed across multiple documents [37].

Documents were uniquely identified by PMCID for biomedical literature, NCTID for clinical trials, and the safety report identifier for FAERS. Scores were calculated independently within each source.

For a drug-ADE pair *t* in document *j*, term frequency was calculated as:

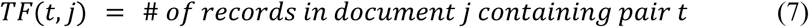

This represents the number of occurrences of the pair within a single document. The source-level term frequency was calculated as:

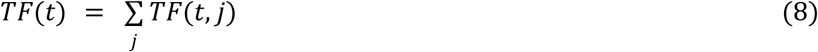

Document frequency was defined as:

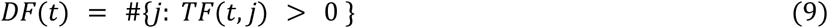

Let *N* represents the total number of unique documents in the source. We therefore calculated TF-DF as:

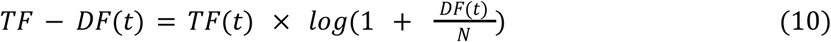

where

- *TF*(*t*) quantifies the total number of observations for a pair,
- *DF*(*t*) measures its distribution across documents,
- *log*(1 + *x*) transformation moderates the contribution of document prevalence.

Source-specific thresholds were used to accommodate differences in corpus size and structure. In the primary analysis, pairs with TF–DF scores at or above the 99th percentile of the source-specific score distribution were designated as high-scoring candidates.

##### Machine Learning Models

We trained five supervised machine learning models to prioritize candidate drug-ADE relationships: Logistic Regression (LR), Random Forest (RF), Gradient Boosting (GB), Support Vector Machine (SVM), and XGBoost. The training set comprised drug–ADE relationships corroborated across multiple sources. Model features (*X*) consisted of source-specific drug-ADE reporting frequencies from FAERS (*F_f_*), biomedical literature (*F_l_*), and clinical trials (*F_c_*):

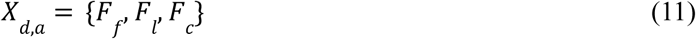

We constructed a gold-standard dataset *Y* curated by licensed pharmacists, where *y_i_* = 1 predicts a valid ADE and *y_i_* = 0 indicates a non-ADE relation.The model was optimized to learn the mapping function *P*(*y* = 1|*X*), representing the probability of a positive label conditional on the observed source-specific features. For each candidate pair, the predicted probability was used as its certainty score (*S*)

#### 2.3.3 Ingredient-based Inference and EHR Integration

##### ADE inference

We used shared active ingredients to expand the representation of drug-ADE relationships across medication products. Let (*D*) denote the set of medication products and (*I*) the set of unique active ingredients. For each medication *d*∈*D*, *I*(*d*)⊆*I* denote its set of ingredients, and *A*(*d*) its set of documented ADE associations.

We first identified single-ingredient medication products:

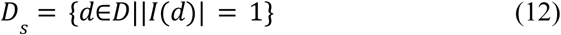

For each ingredient (*i*∈*I*), we identified the set of medication products containing that ingredient:

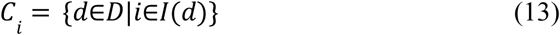

We then performed a set-union expansion to ensure that every medication in the set inherited the documented ADEs from its corresponding single-ingredient reference. The expanded ADE set for medication *d* was defined as:

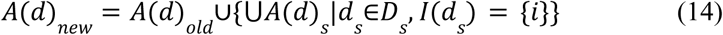

The inferred relationships were represented independently using the relation type *has_inferred_ADE*, which allowed the knowledge expansion without conflating inferred relationships with directly observed or extracted relationships.

##### EHR data Integration

We processed allergy and adverse reaction records from the MGB EHR system [27], [28], [29], mapping recorded medications and adverse reactions to standardized concepts at the product and active-ingredient levels. EHR-derived drug–ADE relationships were incorporated during subsequent knowledge augmentation to extend the relationships established from FAERS, biomedical literature, and clinical trials. These relationships expanded the KG with clinical observations from routine care but were excluded from cross-source corroboration.

### 2.4 Knowledge Visualization through Web-based Interface Development

We developed a web-based interface (video demo is included in the supplementary) to support interactive exploration of the ADE KG. The application used a decoupled client–server architecture. A Python-based RESTful application programming interface (API) communicated with the Neo4j database and translated user requests into Cypher queries over ontology-linked entities. Query results were serialized as JSON and returned to the frontend for interactive rendering.

To improve interpretability, the interface limited displayed ADE-related associations to a specified number of high-ranking relations for each query, ranked by cumulative case counts where these counts were available. Interactive controls enabled users to inspect and compare relationships in the resulting graph.

The interface also supported temporal visualization of ADE-related observations using Chart.js. EHR-derived reporting ratios were transformed into rates per 100,000 cases before visualization.

Temporal trends were displayed as line plots across five-year intervals (Fig. 5b). These visualizations were intended to support exploration of recorded observations and reporting patterns rather than provide population-level estimates of ADE incidence.

### 2.5 Knowledge-enable AI applications to Clinical Care and Research

#### 2.5.1 Knowledge-enabled AI Applications: RAG-based Graph-to-Text Generation with Human-in-the-Loop Prompt Optimization

We validated whether the developed KG can improve LLM-generated, medication-specific ADE summaries using a RAG framework (Fig. 6a) . The experimental design considered three factors: language model (MedGemma or GPT-4o-mini), KG augmentation (with or without KG integrated information), and prompt design (basic or advanced). The prompts are provided in the Supplementary Information.

For each medication, we retrieved its associated ADE subgraph and supplied the resulting structured information to the language model as contextual evidence. In the no-KG conditions, the model generated its summary without the retrieved KG context. The models were prompted to generate medication-specific ADE summaries under the respective basic or advanced instructions.

Prompt development involved two rounds of iterative refinement informed by expert feedback. The advanced prompt was designed to improve the organization and interpretation of ADE information, including the prioritization of relationships based on their evidence characteristics. The 30 medications used in the final evaluation were separate from those used during prompt development, minimizing potential bias from prompt optimization.

We randomly selected 30 medications and generated outputs under all eight experimental conditions. Two domain experts with experience in biology and clinical informatics independently rated each output on comprehensiveness, accuracy, and relevance using a five-point Likert scale, with higher scores indicating better performance. An overall quality score was calculated as the mean of the three criteria.

#### 2.5.2 Knowledge-enabled AI Applications: KG-based ADE Relation Extraction

We evaluated whether KG-derived knowledge could improve drug–ADE relation extraction from clinical narratives. The evaluation included the 2018 National NLP Clinical Challenge (n2c2) ADE dataset, consisting of 505 inpatient notes, and 2,555 outpatient encounter notes collected at Brigham and Women’s Hospital between October 1, 2018, and December 31, 2019 [38], [39].

We implemented a two-stage extraction pipeline using MedGemma, GPT-4o, and GPT-4o-mini. In the first stage, the model identified medication mentions in the clinical text using a predefined extraction prompt. In the second stage, each identified medication was used to retrieve the corresponding ADE subgraph from the KG. The retrieved information was supplied as contextual evidence to the model, which extracted drug–ADE relationships from the original clinical narrative using a structured prompt. The extraction prompts are provided in the Supplementary Information.

We compared KG-supported extraction with the corresponding finetuning approaches without KG-derived context. Performance was assessed against the reference annotations using precision, recall, and F1 score. These metrics quantify the accuracy and completeness of extracted relationships relative to the reference annotations.

### 2.6 Evaluation and Statistical Analysis

#### Evaluation of information extraction and concept normalization

We randomly sampled 80 records, including 40 biomedical abstracts and 40 clinical trial records for independent evaluation by two domain experts. For relation extraction, the reviewers independently annotated drug–ADE relationships directly from the source text while blinded to system output. Inter-annotator agreement was measured using pairwise F1 score by treating one annotator’s relationship set as the reference and the other’s as the predicted set. Disagreements were resolved through consensus to establish a reference annotation set. System-extracted relationships were then compared with this reference set to calculate precision, recall, and F1 score.

For concept normalization, the reviewers independently classified SapBERT-generated UMLS mappings as correct or incorrect. Inter-rater agreement was measured using Cohen’s kappa [40]. Normalization accuracy was calculated as the proportion of mappings judged correct after consensus adjudication.

#### Pharmacist review of drug–ADE relationships

Two licensed pharmacists independently reviewed candidate drug–ADE relationships and classified each as “Yes,” “Maybe,” or “No.” Disagreements were resolved through discussion. To maintain a conservative inclusion standard, only relationships classified as “Yes” were included in the final KG; “Maybe” and “No” relationships were excluded. Pharmacist judgments were also used to establish labels for supervised model development and evaluate computational prioritization.

For supervised prioritization, we assessed discrimination using the area under the receiver operating characteristic curve (AUROC). Precision, recall, and F1 score were used to characterize the trade-off between false-positive and false-negative classifications. Recall was interpreted as the proportion of reference-positive relationships recovered, whereas precision represented the proportion of predicted-positive relationships supported by the reference labels. KG concept coverage and relationship recall were additionally evaluated against the six independent benchmark datasets[30], [31], [32], [33], [34], [35].

#### Statistical analysis of graph-to-text generation

The graph-to-text evaluation followed a balanced 2 × 2 × 2 factorial design involving language model (MedGemma or GPT-4o-mini), KG augmentation (with or without KG), and prompt design (basic or advanced). Each of the 30 medications was evaluated under all eight conditions, and each output was scored independently by two raters.

Inter-rater reliability was quantified using the intraclass correlation coefficient, ICC(2,1), based on a two-way random-effects model for single ratings. Agreement and consistency estimates were calculated overall and within prompt levels. Both raters were included in the subsequent analyses.

For each rating outcome, we fitted a linear mixed-effects model:

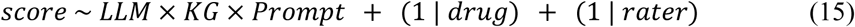

The model included fixed effects for the three experimental factors and their interactions, with random intercepts for drug and rater. Type III F-tests with Satterthwaite degrees of freedom were used to test fixed effects, with sum-to-zero contrasts for the categorical predictors. Estimated marginal means and pairwise contrasts were calculated for KG augmentation, prompt design, and language model, both marginally and stratified by the other factors. Within each outcome, stratified comparisons were adjusted for multiple testing using the Benjamini–Hochberg false discovery rate procedure. Because the basic- and advanced-prompt outputs were generated in separate batches, prompt effects may be partially confounded with batch effects.

All analyses were conducted in R version 4.6.0 using the lme4, lmerTest, and emmeans packages.

## 3. Results

### 3.1 Standardized Extraction and Normalization of Biomedical Evidence from Heterogeneous Sources

We first evaluated the extraction and normalization of drug–ADE relationships from heterogeneous biomedical literature and clinical trial records. LLM-based extraction achieved moderate performance, while ontology-based concept normalization showed consistent accuracy across both data sources, supporting the integration of unstructured biomedical evidence into a standardized knowledge representation.

Manual evaluation of relation extraction (Fig. 2b) yielded an inter-annotator agreement of 0.7222. On biomedical literature abstracts, Qwen3-4B achieved a precision of 0.6410 and a recall of 0.7143, corresponding to an F1-score of 0.6757. Performance was lower on clinical trial records, with a precision of 0.4464, a recall of 0.4237, and an F1-score of 0.4348. These results indicate that extraction performance varied by source, underscoring the importance of assessing information extraction across diverse biomedical data modalities.

For concept normalization, the initial agreement between annotators yielded an F1-score of 0.5952. Discrepancies were resolved through discussion to establish the consensus reference dataset. SapBERT achieved normalization accuracies of 0.8400 for literature abstracts and 0.8243 for clinical trial records (Fig. 2c). The similar performance across these sources supports the use of ontology-based normalization to map extracted concepts to standardized biomedical terminology before information integration.

### 3.2 Evidence-aware Prioritization of Drug–ADE Relationships

We next evaluated strategies for prioritizing candidate drug–ADE relationships across FAERS, biomedical literature, and clinical trial records. Across the three sources, 3,351 medications were identified for harmonization. The number of ADEs associated with individual drugs was highly skewed (Fig. 3b): 1,197 drugs (35.72%) were associated with a single ADE, whereas 0.72% were associated with more than 100 ADEs. Two drugs had the largest numbers of associated ADEs, with 183 relationships each. The source-level analysis showed both overlapping and source-specific drug–ADE relationships across FAERS, biomedical literature, and clinical trial records (Fig. 3c).

Pharmacist review was used to examine agreement between computational prioritization and expert judgments (Fig. 3d). Among the supervised models, the support vector machine (SVM) achieved the highest reported agreement or partial agreement rate, at 73.20% (16,551/22,610), followed by Logistic Regression at 73.04% and XGBoost at 72.92%. In contrast, the TF–DF approach had a disagreement rate of 58.69%. These results show that agreement with pharmacist judgments varied across prioritization methods.

Classification performance also differed according to the definition of a positive relationship (Fig. 3e). Under the inclusive “Yes or Maybe” definition, the Poisson–Gamma empirical Bayesian approach achieved an F1-score of 0.8419 and a recall of 0.9944. TF–DF achieved the highest precision, at 0.7658, but its recall was 0.1173. Among the supervised machine learning models, XGBoost achieved the highest F1-score, at 0.6467. When the stricter “Yes” definition was used, model performance was evaluated against relationships explicitly validated by pharmacists.

Discrimination between positive and negative relationships was further examined using the area under the receiver operating characteristic curve (AUROC; Fig. 3f). XGBoost achieved the highest AUROC under both the inclusive and strict definitions, at 0.9623 and 0.9557, respectively. Gradient Boosting and Random Forest also achieved AUROC values above 0.95 under both definitions, whereas Logistic Regression had lower discrimination performance. Overall, these findings demonstrate that statistical and supervised approaches focus on different aspects of relationship prioritization, with performance dependent on both the method and the definition of a positive relationship.

### 3.3 Integration of Heterogeneous Evidence into a Pharmacovigilance Knowledge Graph

The constructed KG integrated drug, ingredient, ADE, and indication concepts with relationships originated from heterogeneous biomedical data sources (Fig. 4a). The KG contained 11,588 drug nodes, 44,843 ingredient nodes, 3,275 ADE nodes, and 993 indication nodes, connected by more than 2.47 million relationships (Fig. 4b). The principal relationship types included drug–ADE associations *associated_with_ADE* (n=1,683,643) and ingredient–ADE associations (n=721,344). Additional relationships represented drug–ingredient associations *has_ingredient* (n=65,462), potential treatment indications *may_treat* (n=2,046), potential preventive indications *may_prevent* (n=187), and inferred drug–ADE associations *has_inferred_ADE* (n=620).

**Figure 4.**
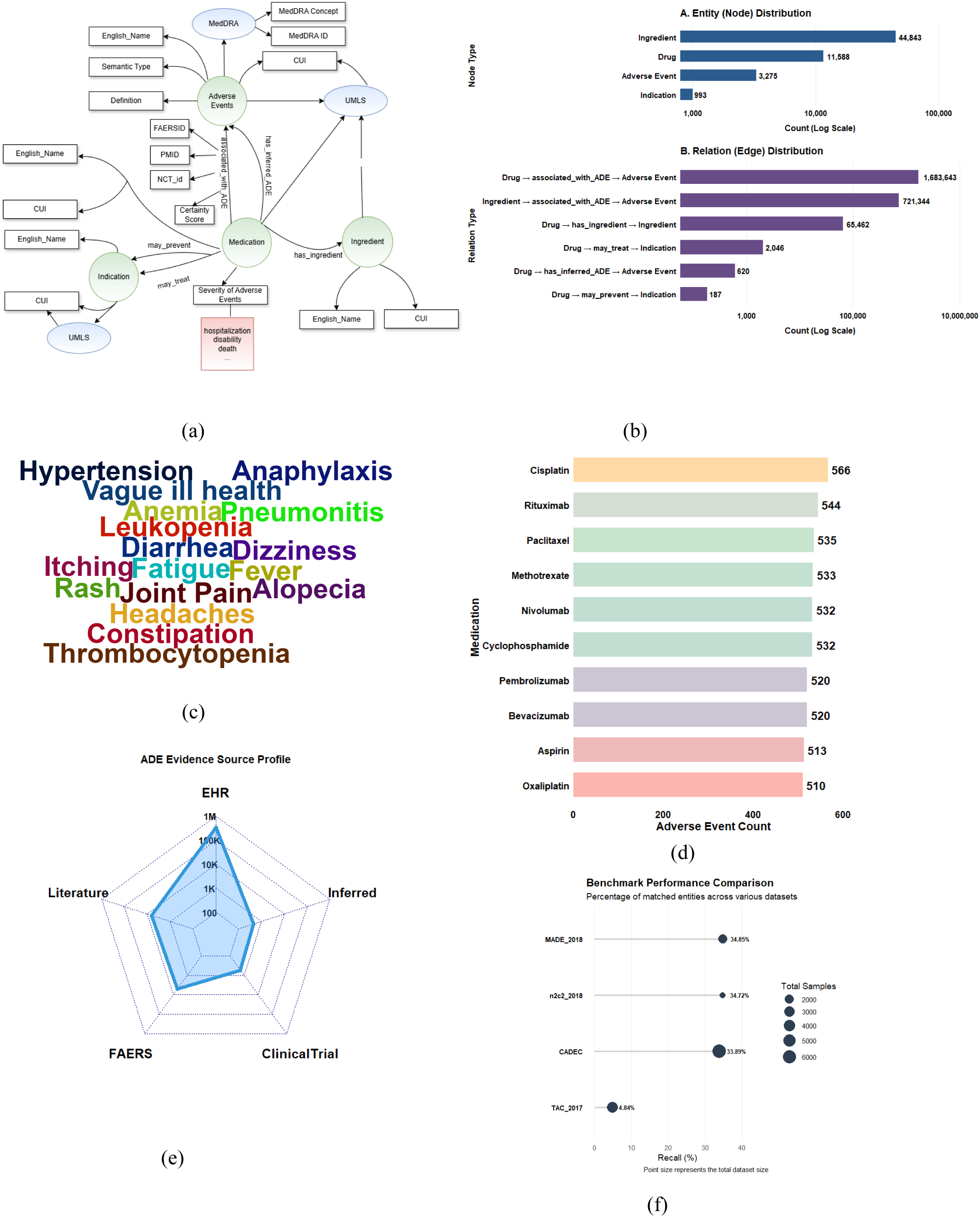
Composition, evidence sources, and benchmark coverage of the pharmacovigilance knowledge graph. a) Overview of the KG architecture and relationships among drugs, ingredients, ADEs, and indications. b) Counts of nodes and edges by entity and relationship type. c) Word cloud visualization showing the most frequent ADEs in the knowledge graph, with font size proportional to frequency. d)Drugs with the largest numbers of associated ADE relationships. e) Distribution of drug–ADE relationships across EHR, biomedical literature, FAERS, clinical trial registries, and inferred associations. f) Recall of ADE relationships against the MADE 2018, n2c2 2018, CADEC, and TAC 2017 benchmarks.

The KG represented a broad range of clinical manifestations (Fig. 4c). Among the most frequently represented ADE concepts were fatigue (n=5,619), diarrhea (n=5,581), and joint pain (n=5,559). The graph also included potentially serious events, including leukopenia, thrombocytopenia, and pneumonitis. Drugs with large numbers of associated ADE relationships included cisplatin (n=566), rituximab (n=544), and paclitaxel (n=535) (Fig. 4d).

The KG integrated drug-ADE relationships originated from large-scale clinical records, biomedical literature, and clinical trials, alongside inferred relationships (Fig. 4e). EHR-derived relationships contributed 2,499,714 drug–ADE relationships, while the KG also included 17,963 relationships from biomedical literature, 13,247 from FAERS, 853 from clinical trial registries, and 638 inferred drug-ADE relationships. We assessed KG coverage against four annotated benchmark datasets (Fig. 4f). The KG achieved recall values of 34.85% on MADE 2018, 34.72% on n2c2 2018, 33.89% on CADEC, and 4.84% on TAC 2017.

Figure 5 demonstrates the practical implementation of ADE KG through a web-based interface, highlighting its capacity to translate complex pharmacological information into actionable insights.

**Figure 5.**
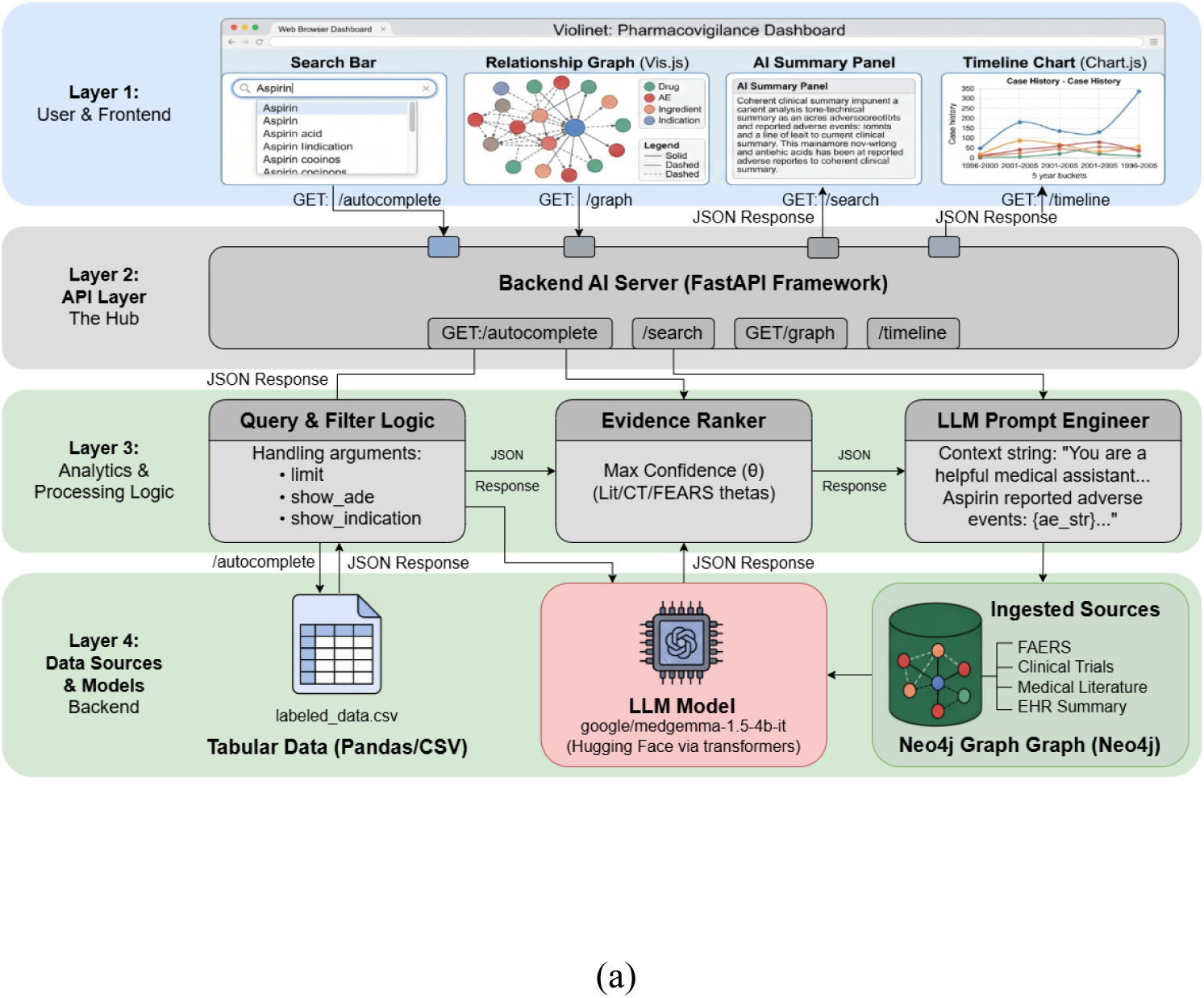
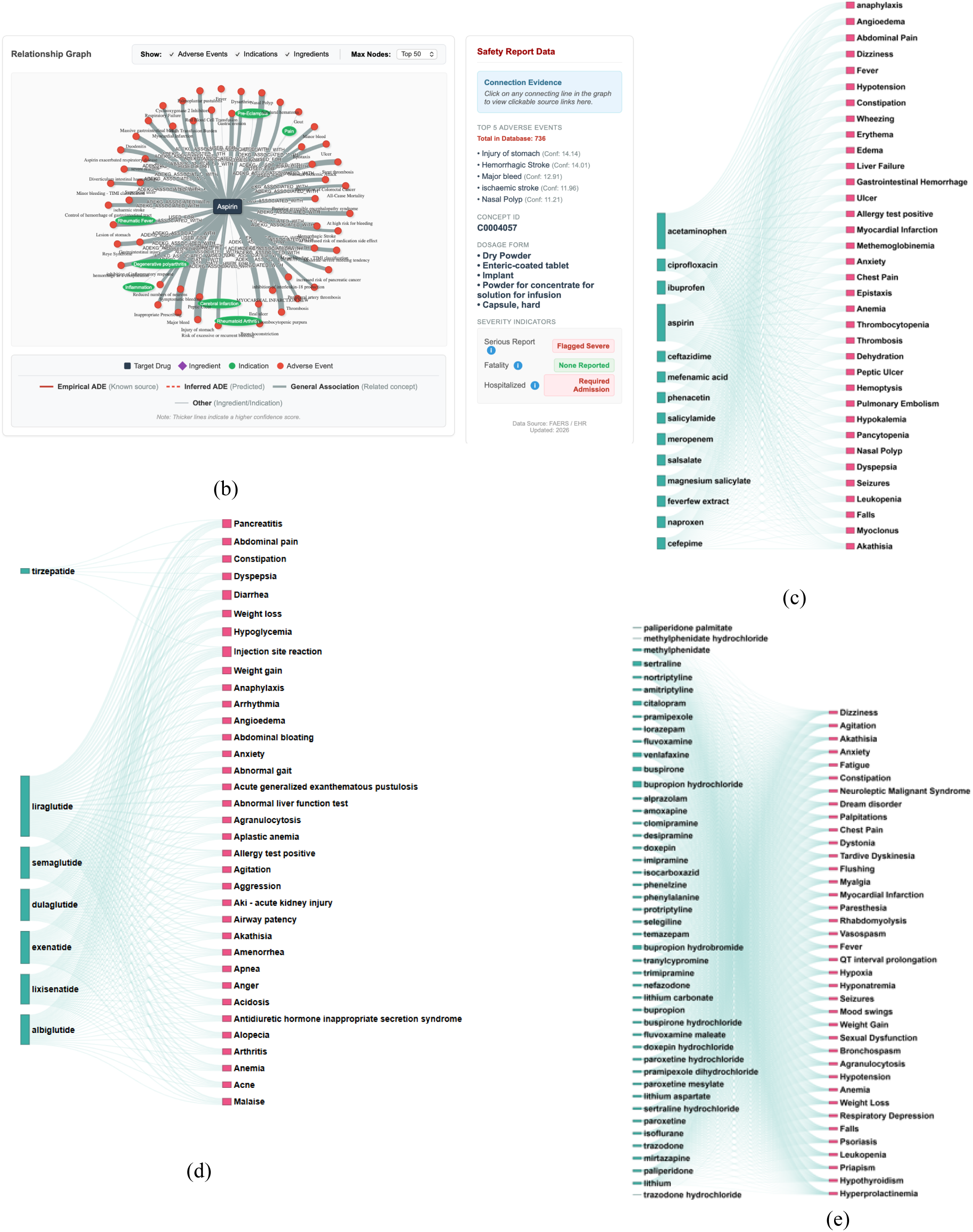
Web-based access to and visualization of the pharmacovigilance knowledge graph. (a) Architecture of the web-based interface and its connection to the KG. (b) Representative interface showing query and visualization functions. (c) Sankey diagram illustrating drug–ADE relationships for drugs indicated for pyrexia (fever). (d) Drug–ADE relationships for GLP-1 receptor agonists. (e) Drug–ADE relationships for antidepressant drugs.

The KG was also made accessible through a web-based interface for querying and visualizing drug–ADE relationships (Fig. 5a,b). Sankey diagrams illustrated the many-to-many structure of associations between drugs and ADEs across selected therapeutic and clinical contexts. For drugs indicated for fever, the visualization included analgesics and antipyretics such as acetaminophen, aspirin, ibuprofen, and naproxen, as well as antibiotics such as ciprofloxacin and cefepime. Associated ADEs ranged from dizziness, fever, and nausea to anaphylaxis, gastrointestinal hemorrhage, and liver failure (Fig. 5c).

Additional examples illustrated the scope of relationships represented for GLP-1 receptor agonists (GLP-1RAs) and antidepressants. For GLP-1 receptor agonists, including tirzepatide, liraglutide, semaglutide, dulaglutide, exenatide, lixisenatide, and albiglutide, the interface displayed associations with events such as abdominal pain, diarrhea, hypoglycemia, pancreatitis, acute kidney injury, and anaphylaxis (Fig. 5d). For antidepressants, including sertraline, citalopram, venlafaxine, bupropion, trazodone, mirtazapine, and amitriptyline, the visualizations included associations with dizziness, fatigue, constipation, QT interval prolongation, seizures, and neuroleptic malignant syndrome (Fig. 5e).

### 3.4 Knowledge Graph Augmentation Improves LLM-based Biomedical Knowledge Synthesis and Clinical Relation Extraction

We validated the downstream utility of the KG in two applications: LLM-based generation of drug–ADE summaries and extraction of drug–ADE relationships from unstructured clinical notes (Fig. 6a,f). For graph-to-text generation, 30 medications were used to compare two language models, KG availability, and prompt configurations. The distribution of supporting evidence varied across drug–ADE pairs and data sources (Fig. 6b). For example, insulin glargine–hypoglycemia and clozapine–neutropenia had reported EHR proportions of 0.391 and 0.356, respectively. Other relationships were more prominently represented in particular sources, including simvastatin–pulmonary toxicity in EHR data (0.186), sertraline–dream disorder in EHR data (0.152), and enoxaparin–major bleeding in biomedical literature and clinical trial metadata (0.188). These examples illustrate variation in the distribution of supporting information across data sources.

**Figure 6.**
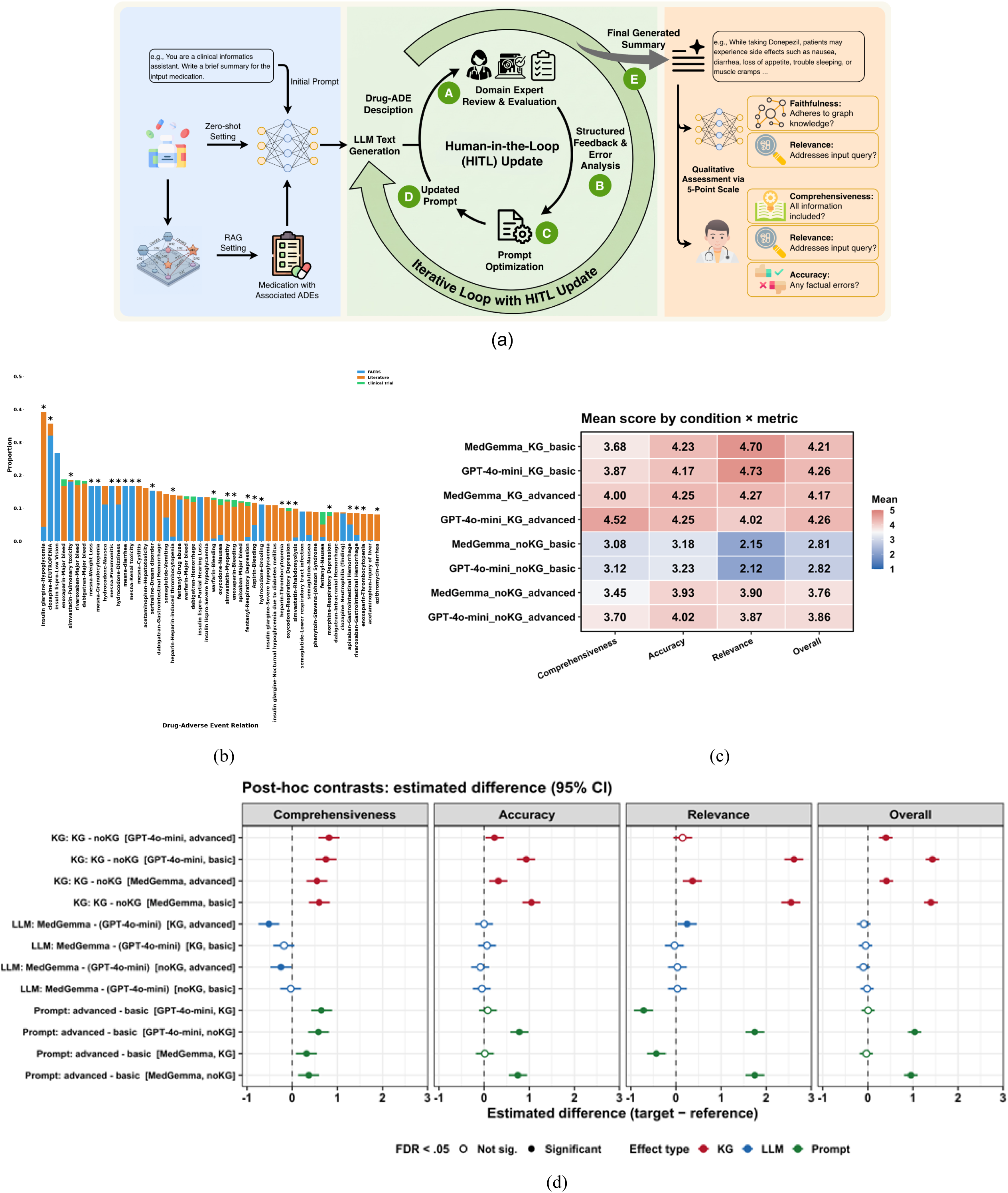
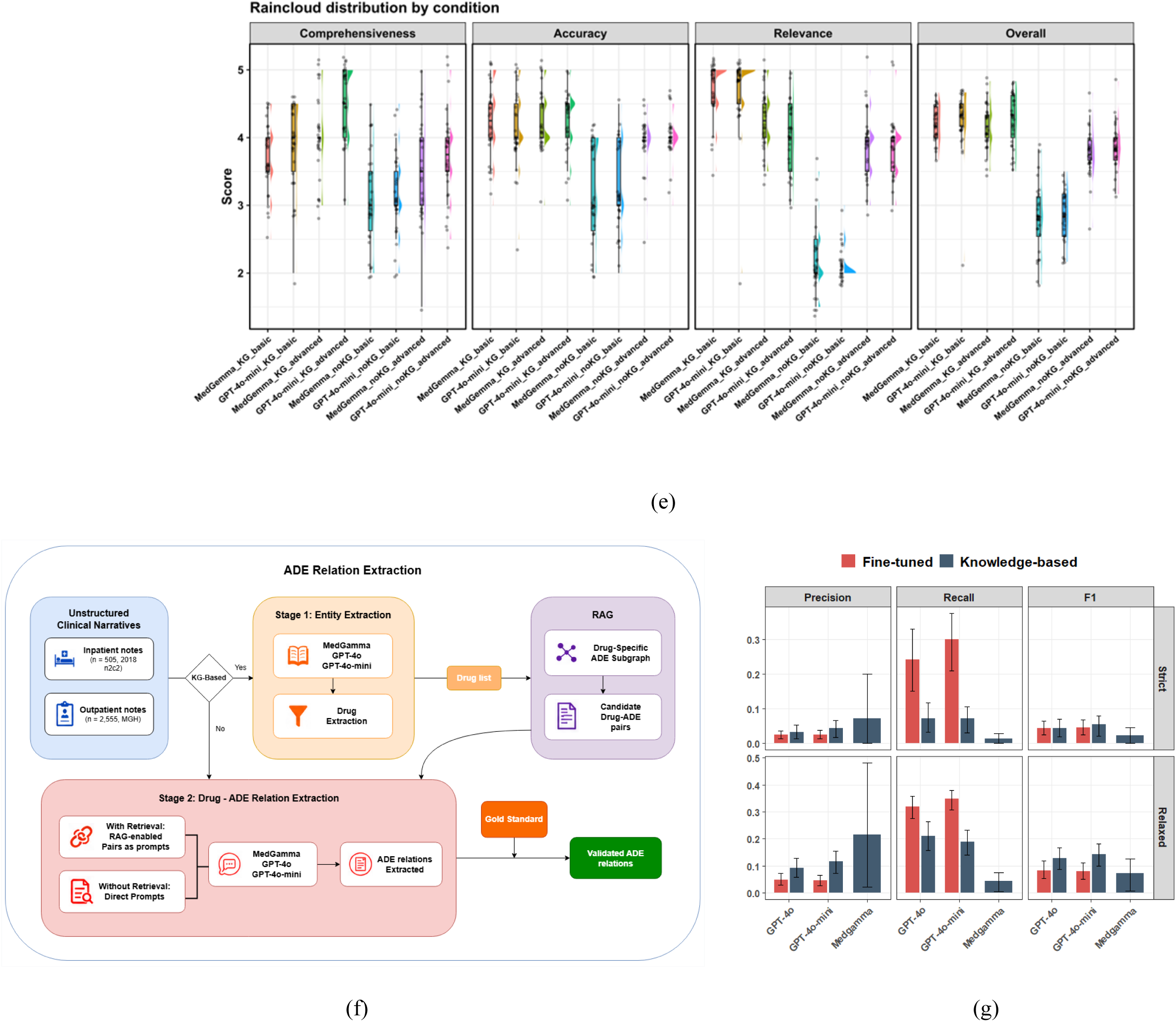
Validation of KG utility via graph-to-text generation and clinical relation extraction. a) Schematic representation of the study framework for graph-to-text generation of ADE descriptions. b) Relative frequency distribution of 50 selected ADEs utilized for graph-to-text generation across heterogeneous data sources; asterisks (*) denote entities documented within electronic health records. c) Heatmap of mean Likert scores across the eight conditions and the three metrics plus Overall. Red = higher, blue = lower. d) Estimated mean differences (95% CI) for each effect family, conditioned on the other two factors. Filled points = FDR < .05. KG (red) is positive and significant in every stratum; the KG benefit is largest under the basic prompt; LLM (blue) is mostly non-significant. e) Raincloud distribution of drug-level (rater-averaged) scores per condition. Flat boxes (e.g., MedGemma / +KG / advanced on Comprehensiveness) indicate a zero inter-quartile range — scores concentrated on a single Likert value. f) Research framework for the extraction of ADE relations from unstructured clinical notes. g) Performance metrics for ADE relation extraction, validating the model’s efficacy in clinical note processing.

Inter-rater reliability for the graph-to-text evaluations was moderate to good across the three rating dimensions. The intraclass correlation coefficient (ICC) was 0.76 (95% CI, 0.70–0.81) for relevance, 0.70 (95% CI, 0.63–0.76) for comprehensiveness, and 0.66 (95% CI, 0.58–0.72) for accuracy. Across the eight combinations of language model, KG availability, and prompt configuration, the integration of KG generally received higher mean ratings than their corresponding conditions without KG information (Fig. 6c). GPT-4o-mini with KG integration and the advanced prompt achieved the highest comprehensiveness score, at 4.52. The highest relevance score was observed with GPT-4o-mini and KG integration using the basic prompt, at 4.73, followed by MedGemma with KG integration and the basic prompt, at 4.70. In contrast, the basic-prompt conditions without KG information had the lowest overall scores, at 2.82 for GPT-4o-mini and 2.81 for MedGemma.

Analysis of estimated mean differences further accounted for the effects of KG availability, language model, and prompt configuration (Fig. 6d). KG integration was associated with a 0.91 increase in overall quality (95% CI, 0.84–0.99) and a 1.42 (95% CI, 1.32-1.53) increase in relevance, the largest estimated effect across the evaluated dimensions. The advanced prompt was associated with an approximately 0.49 increase in overall quality. Differences between language models were smaller, with a statistically significant difference observed only for comprehensiveness. Because the KG-by-prompt interaction was significant, these marginal effects represent averages across the levels of the other factor and should be interpreted in the context of their interaction. Figure 6e shows the distribution of drug-level, rater-averaged scores for each experimental condition. Consistent with the mean scores, the integration of KG exhibited distributions toward higher ratings and generally reduced variability compared with no KG conditions.

We also evaluated KG-based ADE relation extraction from clinical notes (Fig. 6f,g). The instruction-tuned baseline models, GPT-4o and GPT-4o-mini, achieved high recall but low precision under the strict matching criterion, with precision values of approximately 0.025. Incorporating KG information improved precision and F1-score under the evaluated matching criteria. GPT-4o-mini achieved the highest F1-score under both strict and relaxed criteria, with precision increasing by up to 4.8% relative to the fine-tuned baseline models. These results indicate that the KG-based approach can improve the precision of clinical ADE relation extraction while supporting downstream processing of unstructured clinical documentation.

## 4. Discussion

In this study, we developed an AI-empowered evidence-based framework that integrates automated information extraction, semantic normalization, computational prioritization, and knowledge augmentation. The framework applied to transform heterogeneous pharmacovigilance data from spontaneous reporting systems, biomedical literature, clinical trials, and EHR, into a structured, harmonized representation of drug-ADE knowledge graph. Beyond it, the framework also inferred novel drug-ADE relationships based on the shared ingredients. We then leveraged the constructed AI-ready knowledge resource in two downstream AI applications: LLM-based drug–ADE knowledge synthesis and clinical ADE relation extraction. Overall, this study demonstrated how systematic data processing and evidence assessment can support the construction of a structured biomedical knowledge resource and how the developed resource can improve downstream AI applications.

This study has three key contributions compared to existing pharmacovigilance knowledge graphs. First, we developed an AI-ready pharmacovigilance knowledge graph by integrating heterogeneous resources. Our framework explicitly considers the sources and distribution of evidence supporting drug–ADE relationships [21], [22] [23], [24], [25]. Second, we enabled new ADE knowledge discovery through ingredient-based inference, identifying additional drug–ADE relationships across products that share active ingredients. These inferred relationships expand knowledge coverage and retrieval. Third, we incorporated evidence-aware reasoning into KG construction and relationship prioritization. The framework integrates statistical modeling, and supervised prioritization to assess candidate drug–ADE relationships and assigns a certainty score to each included relationship to quantify the degree of supporting evidence. Linking relationships to their source information also allows the KG to be queried in the context of how the underlying knowledge was obtained, supporting a more transparent and informed interpretation of drug–ADE associations. Detailed comparison of our framework with existing systems are shown in Table 2.

**Table 2.** Comparison of our evidence-based framework with other ADE knowledge graph construction approaches.

| Study | How the graph is constructed | Key distinction from our approach |
| --- | --- | --- |
| ADEpedia [13] | Converts drug-label data into RDF, normalizes ADE concepts with biomedical terminologies, and supports review through a semantic web interface. | Our pipeline explicitly prioritizes relationships using statistical and supervised methods and combines evidence from several pharmacovigilance data types. |
| GA-ADE [14] | Builds a graph from literature-derived ADE candidates, identifies core vertices, clusters subgraphs, and estimates drug-event correlations. | Our pipeline involves multiple information sources. |
| DKADE<br>[15] | Uses an ontology to structure drug and adverse-reaction concepts extracted from drug instructions and pharmacopeia, followed by knowledge fusion and inference. | Our pipeline distinguishes cross-source corroboration from separate EHR-based augmentation and validates the curated graph in downstream AI tasks. |

Although this framework was developed for pharmacovigilance, its underlying principles are applicable to heterogeneous data harmonization and integration in other biomedical domains. Our pipeline provides a general approach to transforming diverse biomedical records into structured, AI-ready knowledge. Furthermore, this approach may also support knowledge construction and reasoning in areas such as disease characterization, drug repurposing, precision medicine, and biomedical discovery, where reliable interpretation depends on integrating knowledge across clinical, molecular, and scientific resources.

Several limitations should be considered. First, the KG depends on the coverage and quality of the source data, which vary across medications, ADE types, and clinical contexts. The absence of a relationship from the KG should not be interpreted as evidence that the relationship does not exist [26]. Second, automated extraction and normalization remain imperfect, particularly for complex adverse event descriptions and heterogeneous reporting formats. Third, the current KG does not represent all clinically relevant medication safety knowledge, including contraindications and drug–drug interactions.

## 5. Conclusions

We developed an ontology-driven, evidence-aware knowledge framework that integrates heterogeneous resources into a structured and reusable knowledge base, establishing a generalizable foundation for AI-driven knowledge retrieval, synthesis, and reasoning. We demonstrate how this curated knowledge improves downstream AI performance in knowledge synthesis and clinical ADE detection, highlighting the importance of data curation for reliable AI-driven pharmacovigilance.

## Data Availability

The data generated and analyzed during the current study are not publicly available due to organizational patient privacy policies. Any request to access the Data will need to be reviewed and approved by Mass General Brigham. Researchers will need to provide evidence of IRB approval for their study. For the approved studies, data will be released via a Data Use Agreement.

## Acknowledgments

This article was partially supported by the National Institute of Allergy and Infectious Diseases of the National Institutes of Health under Award Numbers R01AI130460 and U24AI171008, NIH/NLM under Award Number 1R01LM014239 and NIH/NIA under Award Number R01AG080429.

## Competing Interests

The authors declare that there are no competing interests.

## Author Contribution

**C.T.** proposed the research, while **Y.L., L.Z., C.T., J.L., J.Z.,** and **P.H.** designed the study**. Y.L., Z.S., L.W. ,** and **J.P.** collected data, **Z.S.**, **Y.L.**, and **J.Z.** contributed to the information extraction and information normalization. **H.Z., Y.L.,** and **Z.S** developed the methodology of information screening and reasoning, while **H.Z.** and **Y.L.**performed specific experiments. **Y.L., L.Z.,** and **D.L.S.** contributed to the methodology for knowledge graph augmentation, while **Y.L.** performed the corresponding analysis. **F.Y.** implemented the web interface. **C.T.** proposed the idea of graph-to-text generation, while **H.G., J.L.,** and **J.Z.** designed the methodology. **Y.L.** and **F.Y.** conducted the experiments for graph-to-text generation. **Y.L., L.Z.,** and **L.W.** designed the KG-based ADE relation extraction, while **Y.L.** conducted experiments and performed analysis. **S.A.**, **D.L.S.**, **T.C., X.L., J.Z., Z.S., H.L.** evaluated the studies. **M.L.**, **Y.L.**, **D.L.S.**, and **Z.S.** performed statistical analysis. **Y.L., Z.S., M.L., F.Y., H.Z., A.J.,** and **J.Z.** provided the visualization. **J.Z., L.Z., J.L., C.T., Y.H., L.W., Z.S., W.Z.,** and **Y.L.** supervised the study. **L.Z., Y.H., L.C.,** and **D.B.** provided resources. **Y.L., H.Z., H.G., M.L., Z.S.,** and **F.Y.** wrote the manuscript. **J.P., C.T., L.Z., Z.S., Y.H., P.H., J.Z., F.L.,** and **L.W.** critically reviewed the manuscript.

## Notes

### Competing Interest Statement

The authors have declared no competing interest.

### Author Declarations

This study was approved by the Mass General Brigham Institutional Review Board (IRB). The IRB waived the requirement of consent for this study

